# The association between midwifery staffing levels and the experiences of mothers on postnatal wards: cross sectional analysis of routine data

**DOI:** 10.1101/2021.12.14.21267798

**Authors:** Lesley Turner, David Culliford, Jane Ball, Ellen Kitson-Reynolds, Peter Griffiths

**Affiliations:** University of Southampton, School of Health Sciences, Highfield Campus, Southampton SO17 1BJ; NIHR Applied Research Collaboration Wessex, School of Health Sciences, University of Southampton; School of Health Sciences, University of Southampton; Chair of Health Services Research, School of Health Sciences, University of Southampton, National Institute for Health Research Applied Research Centre (Wessex)

**Author notes:** **Corresponding Author** Lesley Turner MSc RN RM.

**Keywords:** Postnatal, Workforce, Midwife, Staffing, Patient experience, Maternity

## Abstract

**Background:** Women have consistently reported lower satisfaction with postnatal care compared with antenatal and labour care. The aim of this research was to examine whether women’s experience of inpatient postnatal care in England is associated with variation in midwifery staffing levels.

**Methods:** Analysis of data from the National Maternity Survey in 2018 including 17,611 women from 129 organisations. This was linked to hospital midwifery staffing numbers from the National Health Service (NHS) Workforce Statistics and the number of births from Hospital Episode Statistics. A two-level logistic regression model was created to examine the association of midwifery staffing levels and experiences in post-natal care.

**Results:** The median full time equivalent midwives per 100 births was 3.55 (interquartile range 3.26 to 3.78). Higher staffing levels were associated with less likelihood of women reporting delay in discharge (adjusted odds ratio [aOR] 0.849, 95% CI 0.753 to 0.959, p=0.008), increased chances of women reporting that staff always helped in a reasonable time aOR1.200 (95% CI 1.052, 1.369, p=0.007) and that they always had the information or explanations they needed aOR 1.150 (95% CI 1.040, 1.271, p=0.006). Women were more likely to report being treated with kindness and understanding with higher staffing, but the difference was small and not statistically significant aOR 1.059 (0.949, 1.181, p=0.306).

**Conclusions:** Negative experiences for women on postnatal wards were more likely to occur in trusts with fewer midwives. Low staffing could be contributing to discharge delays and lack of support and information, which may in turn have implications for longer term outcomes for maternal and infant wellbeing.

**Explanation:** While we recognise that not all gestational parents identify as women; this term was chosen as it has been used in the data source which was accessed for this study and represents most people having maternities.

**Statement of significance (problem):** Women report negative experiences of postnatal care compared with antenatal care and birth. There is a recognised shortage of midwives in maternity services, and this may be impacting on the quality of postnatal care.

**What is already known?:** There is evidence that midwifery staffing levels are associated with birth outcomes but little empirical evidence on the impact of midwifery staffing levels in postnatal care

**What this paper adds?:** This analysis of survey data supports previous findings that increased midwifery staffing is associated with benefits. This is the first study to examine the effects of staffing on women’s experience of postnatal care.

## INTRODUCTION

The State of the World’s Midwifery report estimates that 900,000 more midwives are needed internationally to provide safe care and positive birth experiences ^1^. Investment in midwives has been predicted to substantially reduce maternal and infant mortality^2^ and better workforce planning has been at the forefront of recent policy initiatives ^3-5^.

This study focuses on inpatient postnatal care as women have consistently reported lower satisfaction with postnatal care compared with antenatal and labour care^6-8^. A systematic review of 53 studies on expectations and experiences of inpatient postnatal care concluded that whilst women were generally satisfied with their care, they were sometimes critical of communication, feeding support and a lack of explanations ^9^. In a qualitative study of ten women following caesarean section in Norway, women reported that staff had not fully taken on board their need for rest and adequate pain relief, and were more concerned with aiding breastfeeding than with their post-surgical recovery ^10^. Similar concerns have been raised by researchers in Sweden, who examined feedback from 150 women. They reported that women felt insufficient attention was given to their physical and emotional needs and some felt neglected ^11^. In a survey of 1290 first time mothers relating to care in the first 24 hours after birth, only 41% felt they had all the emotional support they needed, 45% had all the information and advice they needed, and 56% had all their physical care needs met^12^. Fawcett ^13^ conducted a thematic analysis of women’s experience on the postnatal ward. Some women report that hospital staff seemed stressed and overworked, and this impacted on whether they felt able to ask for help^13^.

An association between registered nurse staffing and outcomes for acute inpatients has been found in many cross sectional studies, large longitudinal studies and evaluations of staffing policies^14-16^. Higher skill mix (i.e. total proportion of hours provided by registered nurses) has also been associated with improved outcomes ^17^. There is much less evidence of the impact of staffing in maternity services. This is especially notable in postnatal care, as a recent scoping review found a only a small number of studies, which examined outcomes such as breastfeeding and neonatal readmissions, and none looked at the woman’s postnatal experience in relation to staffing levels^18^.

There has been an increase in caesarean section rates worldwide^19^ which impacts on the complexity of care on postnatal wards and length of stay^20^. The rise in maternal age and obesity also increases the risk profile for some women^21,22^. Women in the UK are now offered discharge from hospital 24 hours after caesarean section if they are recovering well and do not have complications^20^, which alters the acuity case mix of women remaining in postnatal areas. It is important to know whether in-patient services are staffed appropriately for the level of demand and patient acuity, as little evidence currently exists on which to base staffing decisions^23,24^.

In England an annual national survey of maternity care is undertaken which asks all women about experiences of care, including postnatal care. Similarly, midwifery staffing levels are routinely reported for all National Health Service (NHS) care providers, which deliver the majority of maternity care in England. Our research aims to examine whether the quality of postnatal care reported by women is associated with variation in midwifery staffing levels.

The following research question was addressed: Is registered midwife staffing at organisational level associated with variation in women’s experience of inpatient postnatal care, controlling for other factors?

## METHODS

This study is a cross sectional analysis of linked routinely collected datasets in English hospital Trusts. A hospital Trust is an organisation which provides health services in a geographical area in England and manages one or more acute hospitals. Individual patient data on women’s experiences of care has been obtained from the National Maternity Survey 2018 via the UK Data Service^8^. Staffing and workload data was obtained from the NHS Workforce Statistics dataset ^25^ and the Hospital Episode Statistics^26^. The size of the datasets enables detailed statistical analysis of patient and organisation level variables. The use of secondary data is a cost effective way of analysing new questions as data has already been subject to cleaning and quality checks ^27^.

Women were eligible for the annual National Maternity Survey if they had a live birth under the care of a participating hospital Trust during a one-month period and were aged 16 years or over. In 2018, 97.2% of births occurred in an NHS establishment, 0.4% in a non NHS establishment and 2% occurred at home^28^. There is no sampling frame as all women were invited to participate. The survey used postal methodology and women could complete it over the phone if their first language was not English^8^. The Full Time Equivalent (FTE) headcount for midwives was extracted from the NHS Workforce Statistics dataset for February 2018, which corresponds with the same time as the National Maternity survey. Records were linked by the unique hospital Trust code. The number of Full Time Equivalent midwives per 100 births for each hospital Trust was calculated using annual births recorded in the Hospital Episode Statistics.

Data from 129 Trusts and 17,611 women were included in this secondary analysis. The included Trusts represented 98% (129/132) of those offering maternity services. Three Trusts were not included in the survey as they had less than 300 births in the study period. The participating Trusts varied in size, with the mean number of births per Trust reported as 4844 births (range 1122 to 15,500).

The overall response rate was 37% (range of response rates 21% to 61% between Trusts). The majority of women who responded were over 30 years of age (71%). The age profile differed between Trusts e.g. women aged 35 or more ranged from 17% to 57% between Trusts. Data on ethnicity and parity are not available at the individual level but have been obtained at a Trust level from data published online. There is variability in ethnicity of respondents between Trusts, with the lowest proportion of white women in one Trust as 34% and the highest 99%. Overall, 86% per cent of respondents were from a White ethic background, with 8% Asian / Asian British, 3% Black or Black British, 2% mixed ethnicity and 1% Arab or other ethnic group ^8^. Younger women and women from non-white ethnicities were under-represented compared with those giving birth in the same time period^29^.

In the survey, four closed questions asked specifically about the woman’s experience of postnatal care. These relate to whether they experienced a delay in discharge, if they were able to have help within a reasonable time, if they were given the information or explanations they needed, and whether they had been treated with kindness and understanding (Table 1)

**Table 1.**
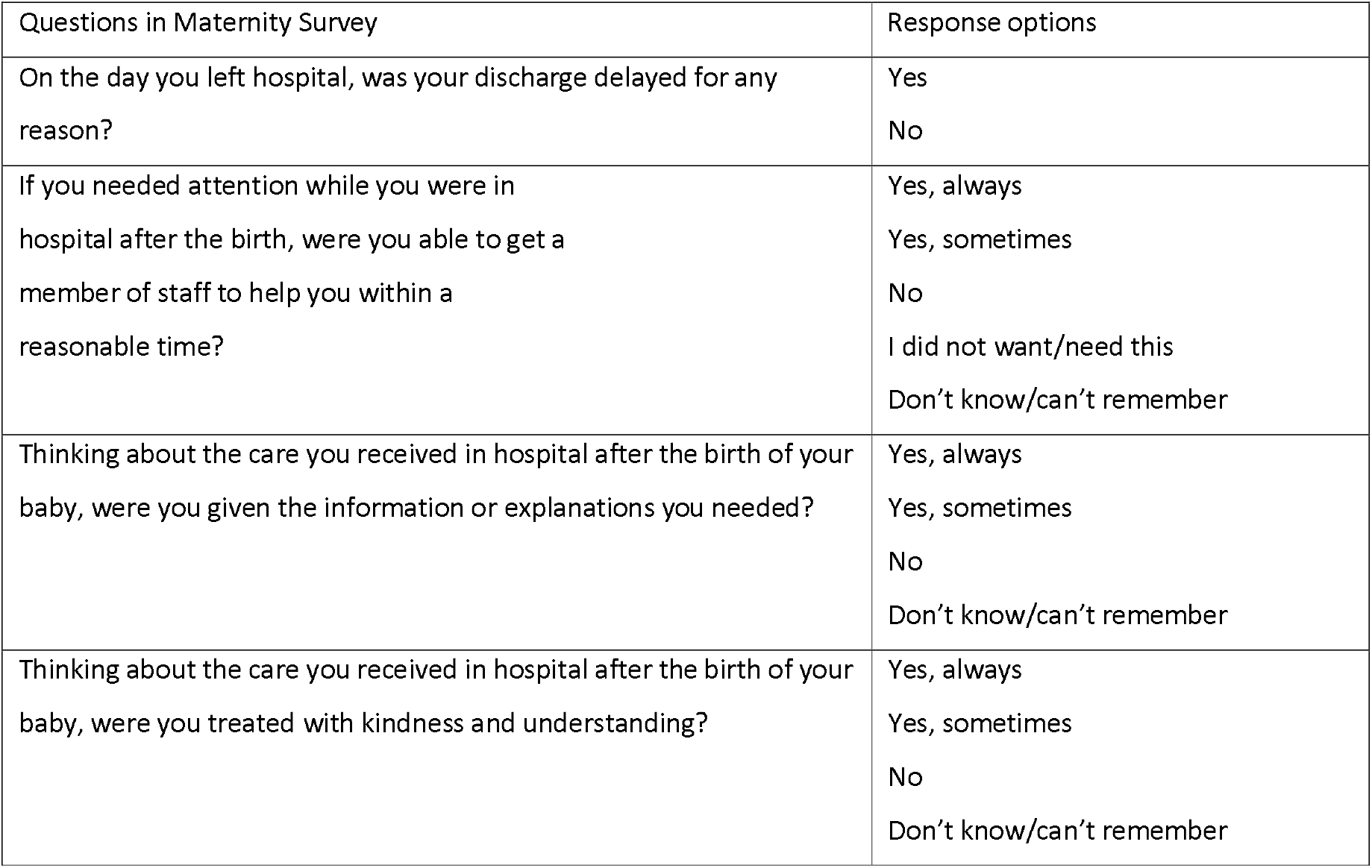
Questions in maternity survey

| Questions in Maternity Survey | Response options |
| --- | --- |
| On the day you left hospital, was your discharge delayed for any reason? | Yes<br>No |
| If you needed attention while you were in hospital after the birth, were you able to get a member of staff to help you within a reasonable time? | Yes, always<br>Yes, sometimes<br>No<br>I did not want/need this<br>Don't know/can't remember |
| Thinking about the care you received in hospital after the birth of your baby, were you given the information or explanations you needed? | Yes, always<br>Yes, sometimes<br>No<br>Don't know/can't remember |
| Thinking about the care you received in hospital after the birth of your baby, were you treated with kindness and understanding? | Yes, always<br>Yes, sometimes<br>No<br>Don't know/can't remember |

These four survey questions were only answered by women who had given birth in hospital, and they specifically relate to postnatal care in that setting. The responses were dichotomised into ‘Yes, Always’ (coded 1) and the alternative which included both ‘Yes, sometimes’ and ‘No’ (coded 0). This grouping was decided in advance of the analysis based on the implied quality standard^30^. A sensitivity analysis was performed by grouping all the ‘Yes’ responses together to examine the effects of this alternative grouping (see Supplementary material). Missing values, don’t know or not applicable responses were removed prior to the analysis, using pairwise deletion. Trust level midwifery staffing levels (as Full Time Equivalent per 100 births) were analysed both as a continuous variable and also divided into tertiles to explore potential non-linear relationships. Three categories were used to ensure sufficient numbers in each category, as the number of Trusts is limited and to aid interpretability.

A two-level multilevel logistic regression model was created using Level-1 (mothers) nested within Level-2 (Trusts). Regression coefficients and adjusted odds ratios (aOR) were calculated for individual predictors as a precursor to fitting a full model. The null model was a two-level random intercept model with no predictors to explore the extent of between-trust variation in the outcomes. Covariates were added to the multilevel models in 3 blocks: i) staffing and number of births per year (Trust level data) ii) age group and type of birth (Individual level data) iii) ethnicity (percentage white) and percentage primiparous respondents (Trust level data).

Akaike’s Information criterion (AIC) and Bayesian information criterion (BIC) goodness of fit data were calculated in order to select models which did not to overfit or underfit the data. Models were selected based on minimising AIC and BIC scores, with lower scores indicating the best fit. Where a difference of less than 2 on the AIC scores was noted, then this was not acted upon as it is not considered to be discriminatory at this level^31^. If AIC and BIC scores disagreed, then priority was given to the model lowest on AIC, and the model lower on BIC was scrutinised and compared for a sensitivity check. Interaction variables of staffing with age, mode of birth and parity were explored to see if this improved model fit using the same method for model selection. To see if results were sensitive to variation in non-responses between Trusts, analyses were repeated including a variable for the Trust response rate and results were scrutinised. The assumptions of the multivariable logistic regression model were examined using guidance by Schreiber-Gregory ^32^. We calculated the number of women who would need to be exposed to a higher staffing level to achieve on additional positive outcome using the reciprocal of the absolute risk difference between staffing tertiles^33^. All data was analysed in Stata 16.1 and coding is presented in the supplementary material.

## RESULTS

For the 129 Trusts studied, the median Full Time Equivalent midwives per 100 births was 3.55 (interquartile range 3.26 to 3.78). This equates to one midwife per 28 births. The distribution of staffing levels shows variation in staffing between Trusts with clustering on the left tail, indicating a positive skew (Figure 1).

**Figure 1:**
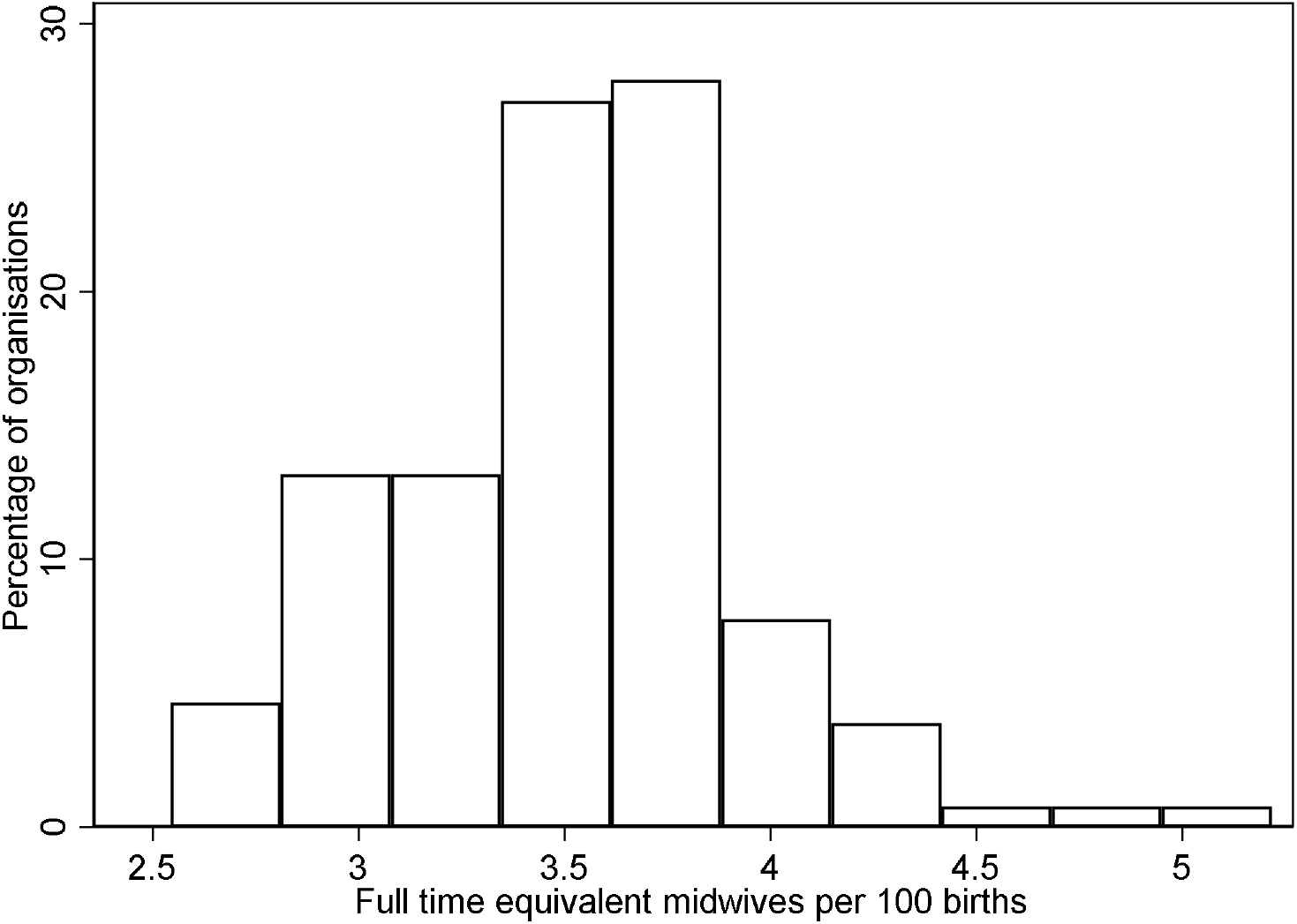
Distribution of Full Time Equivalent midwives per 100 births in the 129 Trusts

The majority of respondents reported that they did not have a delay in discharge (55%) and that staff always helped within a reasonable time (60%), they always had the information or explanations they needed (65%), and were always treated with kindness and understanding (74%). Response categories and frequencies are shown in the Supplementary material. A small proportion of data was missing for each of the four questions, ranging from 466 to 603 respondents (2.6% to 3.4%).

Responses varied by age, type of birth and staffing levels. All the unadjusted rates for categorical variables are presented in Table 2.

**Table 2.** Patient experience outcomes by age, type of birth and staffing levels (% Yes Always)

|  | Delay in discharge | Staff help reasonable time | Information / Explanations | Treated with kindness and understanding |
| --- | --- | --- | --- | --- |
| Responses analysed per variable | 17,050<br>missing 561 | 15,690<br>missing 498<br>Don't Know or NA<br>1423 | 17,027<br>missing 466<br>Don't Know 118 | 16,962<br>missing 603<br>Don't Know 46 |
| Rate - Yes Always as a % of analysed responses | 44.9% | 59.6% | 65.0% | 74.4% |
| Age of mother |  |  |  |  |
| 16-24 | 48.6% | 54.8% | 62.0% | 69.1% |
| 25-29 | 46.1% | 58.8% | 65.3% | 73.8% |
| 30-34 | 44.5% | 59.7% | 63.8% | 74.2% |
| 35+ | 43.7% | 61.0% | 66.8% | 76.0% |
| Type of birth |  |  |  |  |
| Unassisted vaginal delivery | 43.7% | 63.2% | 69.4% | 79.1% |
| Assisted vaginal delivery | 51.9% | 57.0% | 58.0% | 69.5% |
| Planned caesarean | 42.1% | 55.0% | 63.6% | 70.8% |
| Emergency caesarean | 45.2% | 54.4% | 57.9% | 65.8% |
| Staffing levels | Trust level analysis |  |  |  |
| low fte / 100 births<br>fte 2.543 to 3.395 | 47.1% | 57.3% | 63.6% | 73.5% |
| mid fte / 100 births<br>fte 3.396 to 3.706 | 45.6% | 59.7% | 65.0% | 74.0% |
| high fte / 100 births<br>fte 3.707 to 5.217 | 41.6% | 62.1% | 66.8% | 75.9% |

fte = Full Time Equivalent, NA= not applicable

### Predictor variables of age, type of birth, parity, ethnicity, and size of Trust

Older women reported a delay in discharge less frequently (adjusted odds ratio [aOR] 0.808 to 0.892 across older subgroups) and reported more frequently that they were always helped within a reasonable time (aOR 1.206 to 1.473), they always had the information or explanations they needed (aOR 1.177 to 1.339), and had always been treated with kindness and understanding (aOR 1.284 to 1.598). Some variation was also noted in women who had undergone different types of birth. Compared to those having unassisted vaginal birth, those having an assisted vaginal birth were more likely to report delay in discharge (aOR 1.406) and less likely to report that staff always helped within a reasonable time (aOR 0.769), that they were always given the information or explanations they needed (aOR 0.613) or always treated with kindness and understanding so often (aOR 0.604). Similar findings were reported by those who had a caesarean section (Table 3). Full models for each of the outcomes are presented in the Supplementary material.

**Table 3.** Results of multilevel model for predictor variables of age, type of birth, parity, ethnicity and size of Trust

| Multilevel models, nested in Trust. aOR and 95% CI |  |  |  |  |
| --- | --- | --- | --- | --- |
|  | Delay in discharge | Staff help reasonable time | Information / Explanations | Kindness and understanding |
| 16-24 | Reference age group |  |  |  |
| 25-29 | 0.892 (0.784, 1.015)<br>p=0.083 | 1.206 (1.053, 1.382)<br>p=0.007 | 1.177 (1.029, 1.354)<br>p=0.017 | 1.284 (1.114, 1.481)<br>p=0.001 |
| 30-34 | 0.829 (0.733, 0.938)<br>p=0.003 | 1.327 (1.166, 1.511)<br>p<0.001 | 1.139 (1.003, 1.293)<br>p=0.045 | 1.384 (1.209, 1.585)<br>p<0.001 |
| 35+ | 0.808 (0.713, 0.915)<br>p=0.001 | 1.473 (1.291, 1.680)<br>p<0.001 | 1.339 (1.177, 1.524)<br>p<0.001 | 1.598 (1.392, 1.835)<br>p<0.001 |
| Unassisted vaginal birth – reference |  |  |  |  |
| Assisted vaginal birth | 1.406 (1.286, 1.538)<br>P<0.001 | 0.769 (0.700, 0.844)<br>p<0.001 | 0.613 (0.560, 0.672)<br>p<0.001 | 0.604 (0.547, 0.668)<br>p<0.001 |
| Planned caesarean | 0.950 (0.863, 1.046)<br>P=0.298 | 0.673 (0.609, 0.744)<br>p<0.001 | 0.743 (0.673, 0.821)<br>p<0.001 | 0.603 (0.541, 0.671)<br>p<0.001 |
| Emergency caesarean | 1.054 (0.967, 1.149)<br>P=0.230 | 0.688 (0.629, 0.752)<br>p<0.001 | 0.605 (0.554, 0.661)<br>p<0.001 | 0.504 (0.549, 0.554)<br>p<0.001 |
| Trust level characteristics (continuous variables) |  |  |  |  |
| % primiparous | Not in model with best fit | 0.978 (0.967, 0.990)<br>p<0.001 | 0.986 (0.978, 0.995)<br>p=0.003 | 0.983 (0.973, 0.992)<br>p<0.001 |
| % white ethnic group | Not in model with best fit | 1.006 (1.001, 1.011)<br>p=0.023 | 1.003 (0.999, 1.006)<br>p=0.177 | 1.006 (1.001, 1.009)<br>p=0.007 |
| No births per Trust | 1.000 (1.000, 1.000)<br>p=0.536 | 1.000 (1.000, 1.000)<br>p=0.959 | 1.000 (1.000, 1.000)<br>p=0.601 | 1.000 (1.000, 1.000)<br>p=0.296 |

The size of the Trust in terms of the annual number of births did not appear to be associated with variation in any of the reported experiences. The case mix of each Trust suggested that there may be small differences in the experience of cohorts of women in Trusts with more primiparous women (aOR 0.983 to 0.986 for four outcomes) and those with fewer white women (aOR 1.003 to aOR 1.006), however the effect sizes were very small (Table 3).

### Effects of staffing variation

Table 4 shows the association between midwifery staffing and experience for both staffing as a continuous variable and by staffing level tertiles. Full models can be found in the supplementary material.

**Table 4.** Outcomes by staffing levels

|  | Multilevel models, nested in Trust. OR and 95% CI |  |  |  |
| --- | --- | --- | --- | --- |
|  | Delay in discharge | Staff help reasonable time | Information / Explanations | Treated with kindness and understanding |
| FTE midwives per 100 births unadjusted | 0.855 (0.761, 0.961)<br>p=0.008 | 1.246 (1.087, 1.427)<br>p=0.002 | 1.171 (1.059, 1.295)<br>p=0.002 | 1.110 (0.988, 1.247)<br>p=0.080 |
| FTE midwives per 100 births adjusted | 0.849 (0.753, 0.959)<br>p=0.008 | 1.200 (1.052, 1.369)<br>p=0.007 | 1.150 (1.040, 1.271)<br>p=0.006 | 1.059 (0.949, 1.181)<br>p=0.306 |
| Reference group low fte per 100 births 2.543 to 3.395 (Adjusted ORs presented) |  |  |  |  |
| mid fte per 100 births 3.396 to 3.706 | 0.920 (0.815, 1.037)<br>p=0.173 | 1.141 (0.999, 1.303)<br>p=0.051 | 1.089 (0.985, 1.203)<br>p=0.096 | 1.067 (0.956, 1.189)<br>p=0.246 |
| high fte per 100 births 3.707 to 5.217 | 0.789 (0.697, 0.894)<br>p<0.001 | 1.191 (1.037, 1.367)<br>p=0.013 | 1.130 (1.018, 1.255)<br>P=0.022 | 1.080 (0.963, 1.211)<br>p=0.187 |
| Absolute risk difference compared to reference group of low fte per 100 births , predicted from model |  |  |  |  |
| mid fte per 100 births | -2.0%<br>(-5.0%, -0.9%) | 3.1%<br>(0.0%, 6.2%) | 1.9%<br>(0.3%, 4.1%) | 1.2%<br>(-0.8%, 3.2%) |
| high fte per 100 births | -5.7%<br>(-8.8%, -2.7%) | 4.1%<br>(0.9%, 7.3%) | 2.7%<br>(0.4%, 5.1%) | 1.4%<br>(-0.7%, 3.6%) |
| Number of women exposed to provide benefit for 1 woman (calculated as 1/absolute risk difference) |  |  |  |  |
| Highest tertile of staffing vs Lowest tertile | 18<br>(11 to 37) | 24<br>(14 to 111) | 37<br>(20 to 250) | 71 (non significant) |

fte = Full Time Equivalent

In the multi-level model we found that every additional Full Time Equivalent midwife per 100 births was associated with 15% reduction in odds of reporting delay in discharge (aOR 0.849, 95% CI 0.753, 0.959, p=0.008), a 20% increased odds of women reporting that staff always helped in a reasonable time (aOR1.200, 95% CI 1.052, 1.369, p=0.007) and a 15% increased odds of women always having the information or explanations they needed (aOR 1.150, 95% CI 1.040, 1.271, p=0.006). For women being treated with kindness and understanding, the point estimate is in the direction of improved experiences with more staffing, but the relationship was not statistically significant in the adjusted model (aOR 1.059, 95% CI 0.949, 1.181 p=0.306) (Table 4).

Based on these models we estimate that a Trust in the highest staffing tertile would have 5.7% (95% CI 2.7%, 8.8%) fewer women reporting a delay in discharge compared to a Trust with staffing in the lowest tertile. For every 18 women who receive care in these Trusts one fewer would experience a discharge delay in the higher staffed Trust (number needed to be exposed). Trusts with the highest tertile of staffing would have 4.1% (95% CI 0.9%, 7.3%) more women saying that staff always helped in a reasonable time and 2.7% (95% CI 0.4%, 5.1%) more reporting that they had always been given the information or explanations they needed. This equates to one improved outcome in a high staffed Trust for every 24 women or 37 women respectively (Table 4). There appears to be a dose response effect as Trusts with mid-tertile staffing had predicted effects in between the lowest and highest values (Table 4).

### Sensitivity analyses

As Trust response rates varied (from 21% to 61%) we added a variable for the Trust response rate to the model. This led to slightly larger estimates for the effect of staffing but did not substantively alter results. The model with size of organisation, staffing, age group and parity for the outcome of information and explanations had a better fit by the BIC criterion only, however the staffing coefficients were similar to the full model with six predictor coefficients (aOR 1.162 vs aOR 1.150). When alternative dichotomisation was used (‘yes sometimes’ and ‘yes always’ grouped together versus ‘no’) substantive conclusions were generally unchanged, although effect sizes tended to be larger (see Supplementary material for all sensitivity analyses).

Interaction variables improved the model fit for kindness and understanding when staffing levels interacted with age group and the percentage of primiparous women. Some subgroups reported more positive experiences in Trusts with higher staffing, although women aged 25-29 years and Trusts with the smallest proportion of primiparous women had findings in the opposite direction. Introduction of interaction variables did not improve model fit for the other three measures (see Supplementary material).

## DISCUSSION

Midwifery staffing levels varied considerably between hospitals and were associated with variation in a number of patient reported experiences of postnatal care, after adjusting for other variables. Women in Trusts with more midwifery staff were less likely to report they had experienced a delay in discharge. They were more likely to report that staff always helped them in a reasonable time and were always given the information or explanations they needed.

In previous studies, higher midwifery staffing levels have been associated with reductions in postpartum haemorrhage ^34^, reduced need for neonatal resuscitation ^35^, maternal readmission ^36,37^ and increased exclusive breastfeeding rates at discharge^38^. This analysis of survey data supports and expands upon the previous findings that increased midwifery staffing is associated with benefits, by demonstrating differences in important experiences in post-natal care. The effect sizes we observed are relatively small, but large numbers of women are affected, and the adverse experiences may have economic consequences. In the lower staffed Trusts, an estimate of 5.7% more women (1 in 18) reported that their discharge had been delayed. Delay in discharge contributes to bed pressures ^39^ and has negative consequences for the woman’s experience^9^. There is room for improvement as overall 45% of women in the survey reported their discharge had been delayed. In a previous survey, student midwives and postnatal women identified that the postnatal discharge process was rushed and this resulted in poor quality discharge advice^40^. Both a delay in discharge and rushed discharge are unsatisfactory outcomes and have been linked to staffing pressures in postnatal wards.

A higher proportion of staff responding in a timely way and providing information may contribute to a mother’s wellbeing. Psychological health has been recognised as a major public health challenge^41^, with up to one in five women developing mental health problems during pregnancy or in the first year after birth^41^. It is known that women value support, reassurance, and information from health professionals at this time^42^. Previous work has suggested that midwives do not have enough time to talk to women and support them on postnatal wards^9^ and this study provides further evidence that this may sometimes be the case. There may be yet unrecognised consequences of lack of support, which may manifest in readmissions or a decline in breastfeeding rates if not addressed^43,44^. The move to a continuity of carer model in some countries may alter the pattern and experience of care for postnatal women. Such changes require staff to be available at the right time and place to provide this care, and overall staffing levels to facilitate this ^45^.

The increased drive to provide personalised, respectful and compassionate care is seen in many national and international initiatives^46-48^. In this study, three quarters of women felt they were always treated with kindness and understanding. Although the primary analysis did not find a significant association for staffing levels, our sensitivity analyses do provide evidence that that women’s experience of kindness and understanding may be affected by staffing levels. Babaei and Taleghani ^49^ suggested that workload can be a barrier to providing compassionate care. While negative interactions with patients are relatively rare, Bridges, Griffiths ^50^ found that negative interactions are more common with lower staffing levels.

The main limitations of this study are its cross-sectional design and the level at which staffing has been measured. Data from the Maternity Survey has been linked to staffing data at an organisational level, so we do not have an accurate picture of how many midwives were actually deployed in the postnatal ward area. It may be proportional to the total number of midwives, although we cannot be certain of this and recognise there may be variations in how individual Trusts deploy midwives to meet local needs. There may also be registered nurses and non-registered staff in postnatal areas which have not been accounted for, or movement within the organisation during shifts as some midwives may be relocated away from the postnatal ward to meet needs in other areas^44^. Confirmation of these results is needed from studies with more direct measures of postnatal ward staffing. New sources of data from electronic rosters has created the potential to undertake longitudinal studies with exposure to staffing measured at a ward or individual patient level, mirroring studies now being undertaken in general nursing^51^.

We did not consider staffing by other professional groups in this analysis. It is conceivable that other staff groups, such as doctors, adult nurses, neonatal nurses and nursery nurses contribute to women’s experiences in the postnatal period. Although this survey data records women’s perceptions of delays in discharge actual delays have not been empirically demonstrated, and this could be the focus of future research.

This was a large national study including 129 Trusts and over 17,000 women which aids the generalisability of findings. Although the response rate of 37% is fairly typical of similar surveys this does raise questions about the experience and views of non-responders. The study respondents differed from the target population, for example, there were fewer younger mothers and women from non-white ethnic groups ^29^. However, the sensitivity analyses of response rate, did not suggest a bias arising from variation in response rates between Trusts.

This analysis of the Maternity Survey in the UK adds to the body of evidence examining staffing and outcomes in maternity care. We found that that variation in midwifery staffing at organisational level is associated with variation in women’s experiences of postnatal care. Low staffing levels were linked to higher levels of adverse experiences that could have important consequences in terms of hospital resource use and maternal wellbeing. While we cannot assume the relationship is causal, nonetheless it seems plausible and this is worthy of further exploration.

## Data Availability

Data obtained from the UK Data Service/UK Data Archive, including subsets and derived data, cannot be submitted to journals alongside publications as this would be a breach of the End User Licence (EUL) as in https://www.ukdataservice.ac.uk/help/faq/publication.aspx#Journals

https://www.ukdataservice.ac.uk/

## Notes

### Competing Interest Statement

The authors have declared no competing interest.

### Funding Statement

Peter Griffiths receives support from a Senior Investigator award made by the National Institute for Health Research and the National Institute for Health Research Applied Research Centre (Wessex).
This research was part funded by the National Institute for Health Researchs Health Services & Delivery Research programme (Award ID : NIHR128056)
The views expressed are those of the author(s) and not necessarily those of the National Institute for Health Research, the Department of Health and Social Care, arms-length bodies or other government departments.

### Author Declarations

This study involves only openly available data which can be obtained from the UK Dataservice upon request

